# AI Learning for Pediatric Right Ventricular Assessment: Development and Validation Across Multiple Centers

**DOI:** 10.1101/2025.03.14.25323989

**Authors:** Charitha Reddy, Yi Yan, Min Qiu, Yi Tang, Bo Jin, Zhi Han, Yuhang Li, Sihan Zhou, Qiming Tang, Huan Xiao, Shu Yang, Qigui Wen, Lan-Ping Wu, Li-Jun Fu, Ze-Yu Jing, Yi-Jia Yang, Yu-Qi Zhang, Naoto Ozawa, Takumi Ichikawa, Ellen Ling, Ronald J. Wong, Ivana Marić, Nima Aghaeepour, Brice Gaudilliere, Martin S. Angst, Karl G. Sylvester, Harvey J. Cohen, Gary L. Darmstadt, David K. Stevenson, Henry Chubb, Scott Ceresnak, Animesh Tandon, Doff B McElhinney, Hao Zhang, Xuefeng B. Ling

## Abstract

**Background:** Congenital and acquired heart disease affects ∼1% of children globally, with right ventricular (RV) dysfunction being a common and complex issue due to conditions like congenital heart disease (CHD), pulmonary hypertension (PH), and prematurity. Accurate RV assessment is challenging due to its unique geometry, interventricular interactions, and morphological variability in pediatric patients. Fractional area change (FAC), a key echocardiographic measure, correlates strongly with disease severity, aiding in timely intervention and prognosis. AI learning shows the potential to automate and standardize RV assessments, overcoming traditional limitations and improving early diagnosis and management of pediatric cardiovascular disorders.

**Methods:** Using 24,984 echocardiograms from 3,993 pediatric patients across four tertiary care centers (one in North America, three in Asia), we developed and validated an AI framework for automated RV assessment. The framework employs multi-task learning to perform ventricular segmentation, beat-by-beat quantification of RV FAC, and identification of cardiac abnormalities like PH. It was also extended to enhance left ventricular (LV) functional assessment.

**Findings:** Our AI system achieved Dice similarity coefficients of 0.86 (apical-four-chamber, A4C) and 0.88 (parasternal-short-axis, PSAX) for RV segmentation, matching expert annotations. It demonstrated robust RV functional assessment, with AUCs of 0.95 (U.S. cohort) and 0.97 (Asian cohort). For PH classification, diagnostic accuracies were 0.95 (U.S.) and 0.94 (Asian), confirming consistent performance across populations. When extended to LV assessment, the framework significantly improved LV ejection fraction (EF) prediction in both U.S. and Asian cohorts.

**Interpretation:** This validated AI framework enables reliable, automated ventricular function analysis, matching expert-level performance. By enhancing clinical workflows and standardizing pediatric cardiac assessments, it has the potential to improve care management for pediatric cardiovascular disorders, particularly in resource-limited settings.

**Funding:** This work was supported by the U.S. NIH 1R41HL160362-01 to XBL and K23HL150279 to AT.

## Introduction

Congenital and acquired heart disease impacts the pediatric population worldwide, affecting approximately 1% of children globally. Right ventricular (RV) dysfunction is a prevalent form of heart disease in pediatric patients, often arising from conditions such as prematurity, post-surgical effects of congenital heart disease (CHD), functioning as the systemic ventricle, and idiopathic pulmonary hypertension (PH). These patients face significant risks, including RV failure, decreased quality of life, potential need for transplantation, and increased mortality. Accurate assessment of RV function is crucial for diagnosing and managing cardiovascular conditions but is challenging due to the complex RV geometry.

RV dysfunction primarily results from pressure and volume overloads and has unique pathophysiological characteristics. The RV may tolerate changes for extended periods before resulting in symptomatic failure. Furthermore, RV dysfunction frequently leads to LV dysfunction due to strong interventricular interactions. These differences, along with the irregular shape and orientation of the RV, complicate imaging and assessment, requiring advanced imaging techniques, often involving multiple modalities. ^1^ Abnormal RV function is typically defined as a deviation from normal physiological function, often characterized by reduced ability of the right ventricle to contract or relax effectively. ^2^ Precise evaluation of RV size and function is essential for diagnosing, managing, and predicting outcomes in pediatric cardiac conditions. Echocardiography, as a non-invasive and accessible tool, is the first-line imaging technique for monitoring RV function. ^1^ However, traditional RV functional measures face limitations in pediatric populations due to significant variability in RV morphology. Among systolic parameters, fractional area change (FAC) has demonstrated stronger correlations with disease severity in advanced heart failure patients and a closer relationship with RV ejection fraction (EF) as measured by cardiovascular magnetic resonance (CMR), suggesting its reliability in assessing pediatric RV function. ^3,4^ An FAC <35% is considered abnormal by the guideline^5^. Accurate FAC assessment can guide timely interventions, improve prognosis, and enhance long-term outcomes by enabling better monitoring of RV function over time.

Echocardiography is a non-invasive, widely accessible, and essential first-line tool for routine follow-up of various pediatric cardiac conditions affecting the RV. ^1^ However, assessing RV function in pediatric heart disease remains challenging due to significant variability in RV morphology and physiology. Advances in AI-driven echocardiography, particularly deep learning, hold promise for enhancing cardiac function assessment.^6^ Recent innovations, like EchoNet-Dynamic^7^, have shown the utility of video-based deep learning algorithms for adult LV segmentation and EF estimation. Building on this progress, Reddy et al. developed EchoNet-Peds,^8^ an AI model for pediatric echocardiography that automates LV segmentation and EF calculations. Beside LV segmentation methods, ^9-11^ other deep learning applications include automated quantification^12^ of LV structure and function, as well as novel methods for estimating intraventricular hemodynamic parameters on a beat-to-beat basis ^13^. However, most deep learning studies focus on LV assessment, with comparatively fewer models dedicated to the RV, mainly addressing segmentation tasks. ^14,15^ While some recent advancements have emerged, such as a model estimating RV EF from echocardiographic images PH patients ^16^, significant gaps remain in applying AI to pediatric RV functional assessment.

Currently, no existing echocardiogram-based AI tools are capable of providing beat-by-beat predictions of pediatric right ventricular fractional area change (RV FAC) or delivering a comprehensive assessment of pediatric RV function. This study addresses this critical gap by leveraging advanced deep learning algorithms applied to echocardiographic data, aiming to enhance the precision, reliability, and clinical utility of RV functional assessment. Validation across multi-center cohorts, encompassing diverse ethnicities such as American and Asian populations, demonstrates minimal racial variability in the tool’s performance. This underscores the broad applicability and generalizability of our AI-driven solution, making it a promising candidate for widespread adoption in pediatric cardiology practice.

## Methods

### Study Population and Sample Collection

This study utilized data from the Pediatric Ventricle Function Assessment Study (NCT06739057), encompassing 3,993 pediatric patients across four tertiary care centers (one in North America and three in Asia), with 24,984 echocardiographic studies (Figure 1, Table S1). Additional patient enrolment information is described in Supplementary Materials. Echocardiograms were screened for quality based on image clarity, completeness of ventricular views, and clinical data availability. Figure S1 presents a Venn diagram summarizing the overlap among the four studies. Notably, 486 patients were common across all studies, while 525, 1088,94, and 311 patients were unique to Studies 1, 2, 3, and 4, respectively. These findings underscore both the shared and distinctive patient populations, highlighting the potential for both integrative analyses and study-specific insights.

**Figure 1.**
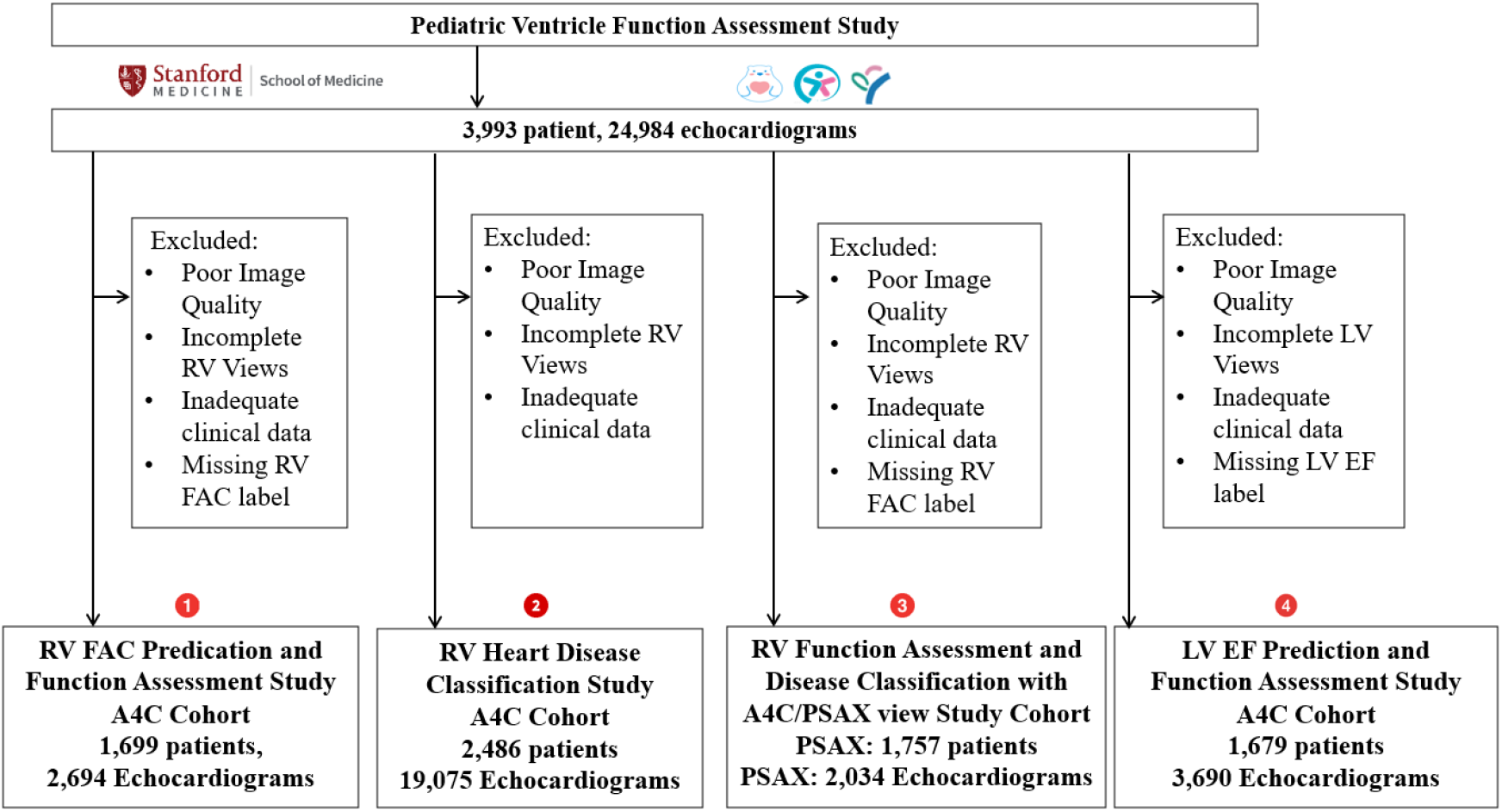
Consort diagram of patient enrollment.

### Study Design

Four cohorts were established for targeted analyses, as shown in Figure 1:

**Study 1. RV FAC prediction cohort** included 1,699 patients and 2,694 echocardiograms for quantitative RV systolic function assessment through FAC. Exclusion criteria included poor image quality, incomplete RV views, inadequate clinical documentation, missing RV FAC label.

**Study 2. RV disease classification cohort** comprised 2,486 patients with 19,075 echocardiograms, dedicated to detecting and classifying RV-related cardiac conditions. Exclusion criteria included poor image quality, incomplete RV views, and inadequate clinical documentation.

**Study 3. RV function assessment and disease classification [A4C and parasternal short-axis (PSAX)] cohorts** included 2,304 PSAX videos from 1,757 patients. Both A4C and PSAX videos were analyzed to evaluate RV function^17,18^ Exclusion criteria included poor image quality, incomplete RV views, inadequate clinical documentation, and missing RV FAC labels.

**Study 4. Exploration of the RV AI learning framework to assess LV. LV EF prediction cohort** included 1,679 patients and 3,690 echocardiograms to apply the RV AI learning framework for LV EF assessment. Exclusion criteria included poor image quality, incomplete LV views, inadequate clinical documentation, and missing LV EF labels.

### U^2^-Net based RV segmentation

The model design and training were implemented in Python 3.10.9 using the PyTorch 2.3.1 deep learning library, which offers a robust framework for building and training neural networks. To achieve automated segmentation of RV regions, a U^2-^Net-based deep learning framework^19^ was employed. Additional details are provided in the supplementary materials.

### RV FAC prediction and function assessment

This study utilized the RV FAC values calculated by on-site cardiologists, as documented in the finalized clinical reports, as the gold standard (ground truth) for training and benchmarking our AI learning algorithms. Using segmentation models, FAC was calculated from area changes across frames. Figure 2 illustrates our RV AI workflow to predict FAC and allow beat-by-beat segmentation (Figure S2) assessment of RV function using single-center and multi-center echocardiograms (Table S2). Additional details are provided in the supplementary materials.

**Figure 2.**
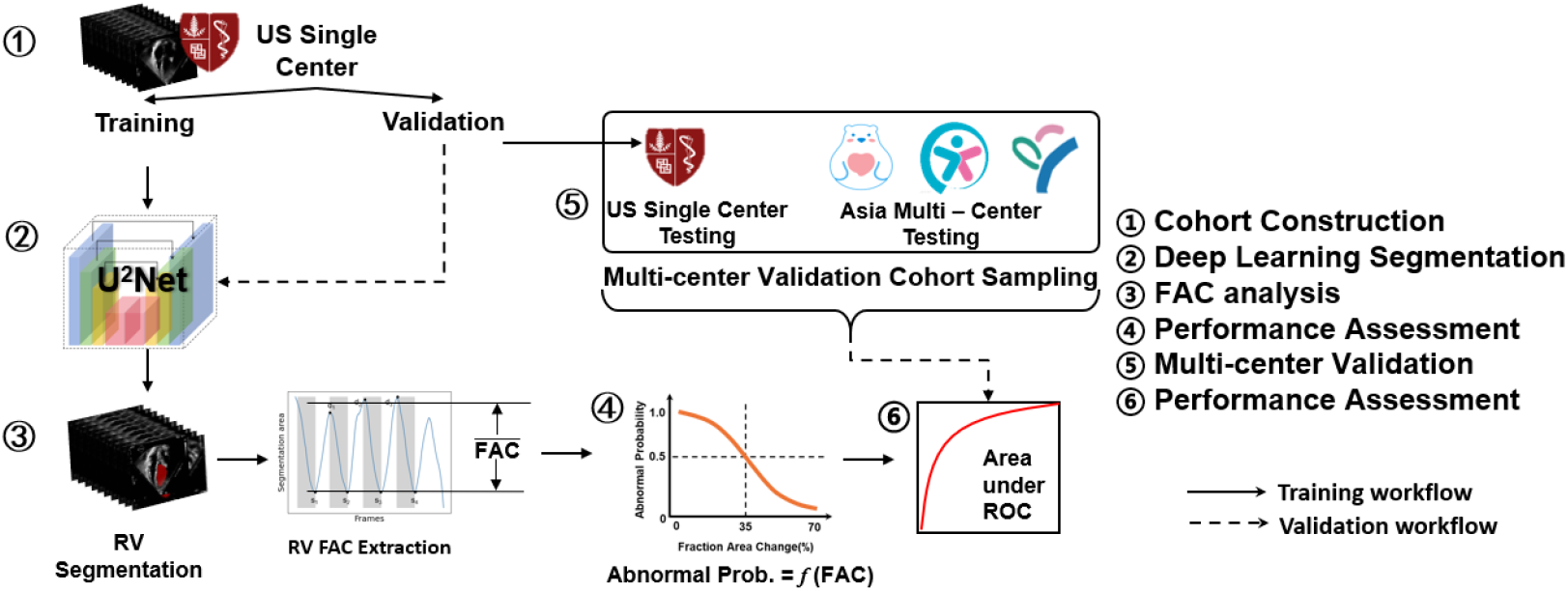
Multi-center RV AI learning workflow to predict FAC and allow automated assessment of RV function.

### RV based CSN modelling for pulmonary hypertension (PH) identification

Building on the above RV segmentation, we further developed Channel Separated Convolutions Network (CSNs)^20^ classification models (Figure 3) to identify PH using multi-center pediatric RV A4C echocardiograms (Table S3). The parasternal-short-axis (PSAX) view is often used alongside the A4C view^21^ to provide a more comprehensive evaluation. In this study, we investigated the combined analysis of A4C and PSAX views (Table S4, Figure S3 and S4) to assess RV function and enable automated identification of pulmonary hypertension (PH). Additional details are provided in the supplementary materials.

**Figure 3.**
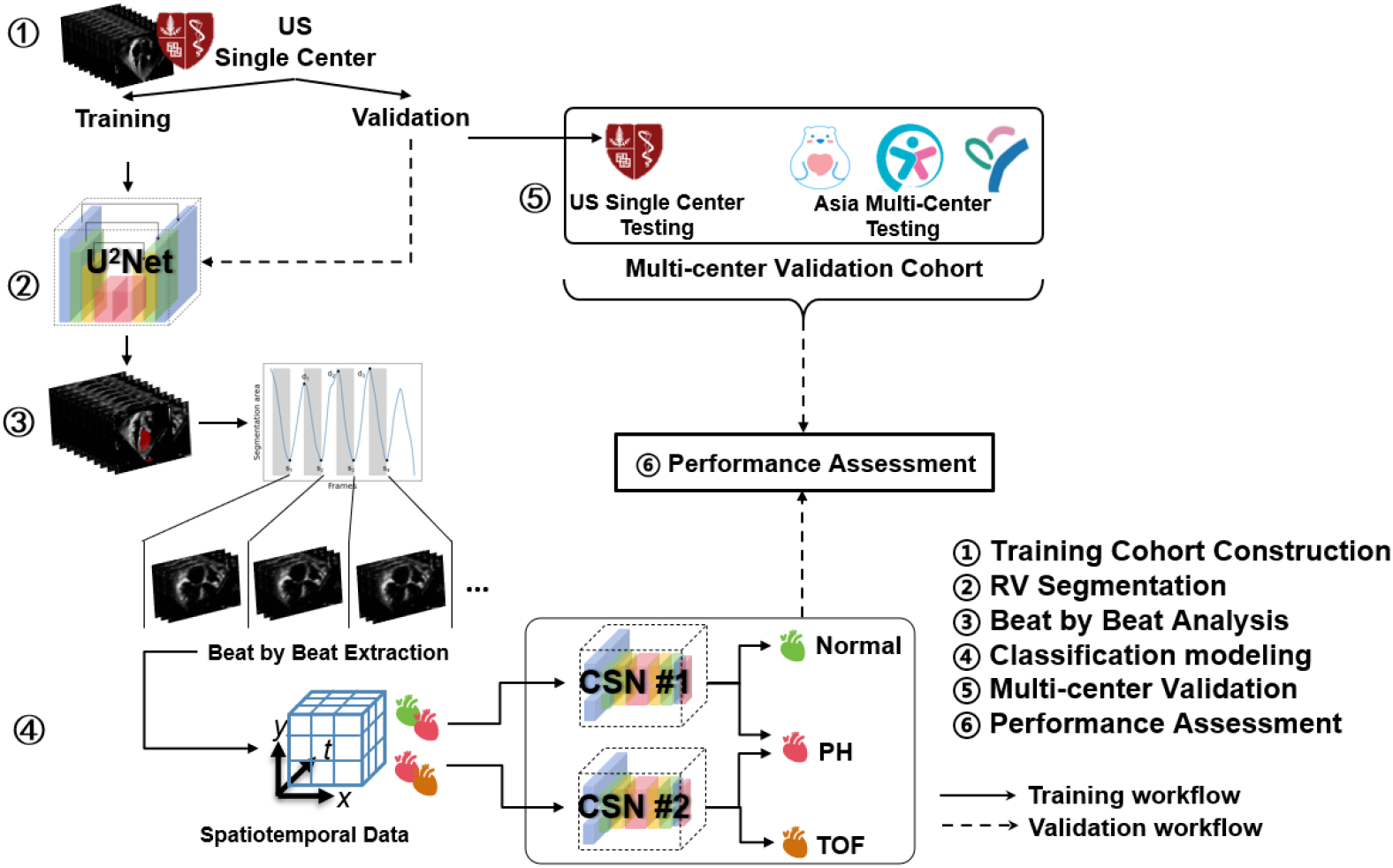
Multi-center RV AI learning workflow to identify PH from other conditions. PH: pulmonary hypertension. TOF: Tetralogy of Fallot. CSN: channel-separated-convolution-network.

### Exploration of the RV AI learning framework to assess LV

This study utilized the LV EF values calculated by on-site cardiologists, as documented in the finalized clinical reports, as the gold standard (ground truth) for training and benchmarking our AI learning algorithms. We applied RV AI learning framework for LV frame-level segmentation and beat-by-beat learning (Figure S5), and EF prediction using echocardiogram videos from a multi-center LV dataset (Table S3). The training and testing groups were older (median 9.5 years), while the Asia cohort skewed younger (median 2.4 years). The U^2^-Net model is applied to perform LV segmentation, followed by LAC (LV 2D Area Change) extraction from the segmented echocardiograms. The extracted LAC, along with patient factors such as age, sex, and heart rate, is used to estimate EF. EF values showed slight regional variation, with an average EF of 64.6% (SD 5.6) in U.S. and 67.9% (SD 3.1) in Asia. Figure S6 illustrates the LV function assessment pipeline on pediatric echocardiograms.

### Statistical Analysis

Results are presented as mean ± SD. FAC estimation accuracy was evaluated using mean absolute error (MAE), root mean squared error (RMSE), and Pearson correlation coefficient (R). Segmentation accuracy was assessed with Dice similarity coefficients.^22^ Diagnostic performance metrics included AUC, sensitivity, and specificity for RV function assessment, as well as accuracy, sensitivity, and specificity for PH/normal and PH/TOF (Tetralogy of Fallot) classifications.

## Results

### Study population

The study cohort comprised pediatric patients from two sources: a single center in the United States (1,763 patients) and multiple centers in Asia (2,230 patients), as detailed in Table S1. A total of 24,984 echocardiogram videos were collected, including 13,369 from U.S. and 11,615 from Asia, with demographic and clinical differences between the cohorts that may impact the results’ generalizability. The median age was 9.6 years (IQR 5–14) for the cohort and 3.7 years (IQR 0.9–4.7) for the Asian cohort. Patients were categorized into four diagnostic groups: normal, PH, TOF, and others. Most patients were classified as normal (1,681 in U.S., 1,531 in Asia), with smaller numbers diagnosed with PH (41 in U.S., 418 in Asia) and TOF (41 in U.S., 281 in Asia).

### RV FAC prediction and RV function assessment

Figure 4 illustrates the model’s performance for FAC prediction and RV function assessment based on the predicted FACs across two cohorts. In the U.S. cohort (Figure 4A), the Dice coefficients for RV segmentation were 0.89 for training and 0.86 for validation. The model achieved R 0.79 for RV FAC prediction (Figure 4B), and AUC (Figure 4C) 0.97 in training and 0.95 in validation for function assessment, demonstrating strong performance. Similarly, in the Asian cohort (Figure 4D, 4E), the model exhibited robust accuracy, with R 0.84, MAE 4.35, and RMSE 6.08 for RV FAC prediction, and AUC 0.95 for RV function assessment.

**Figure 4.**
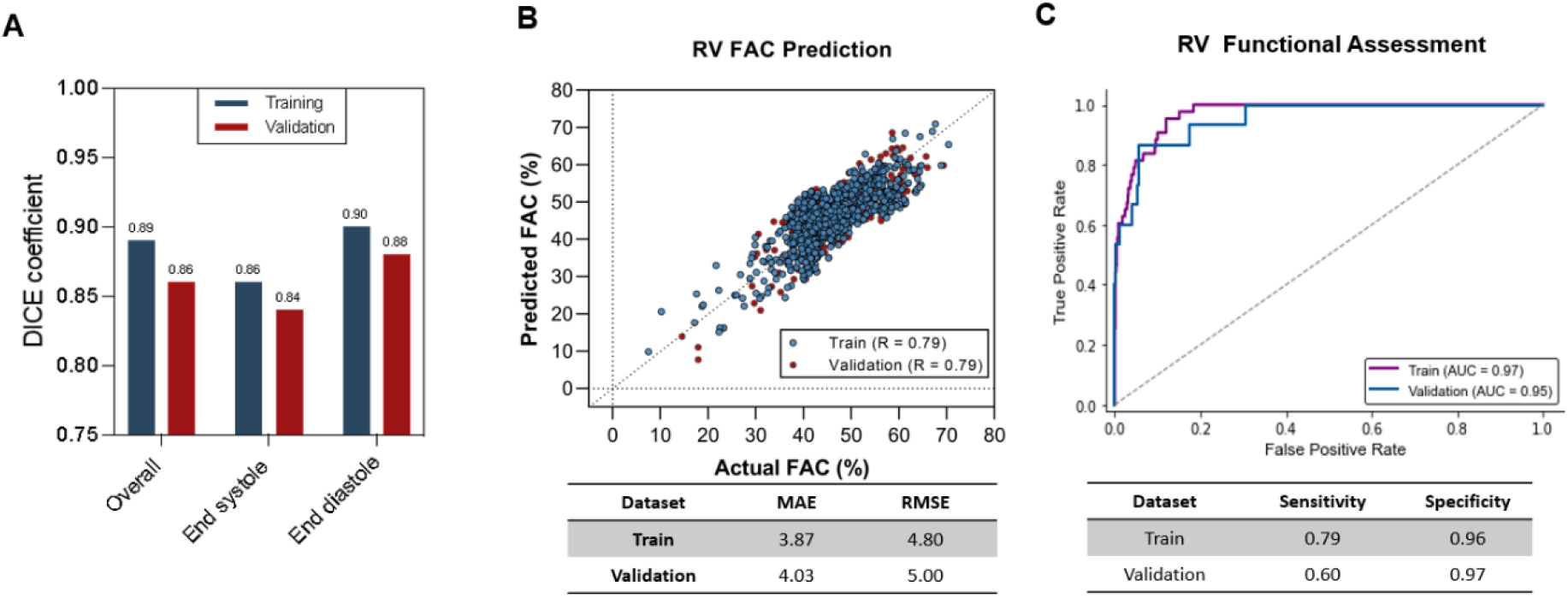
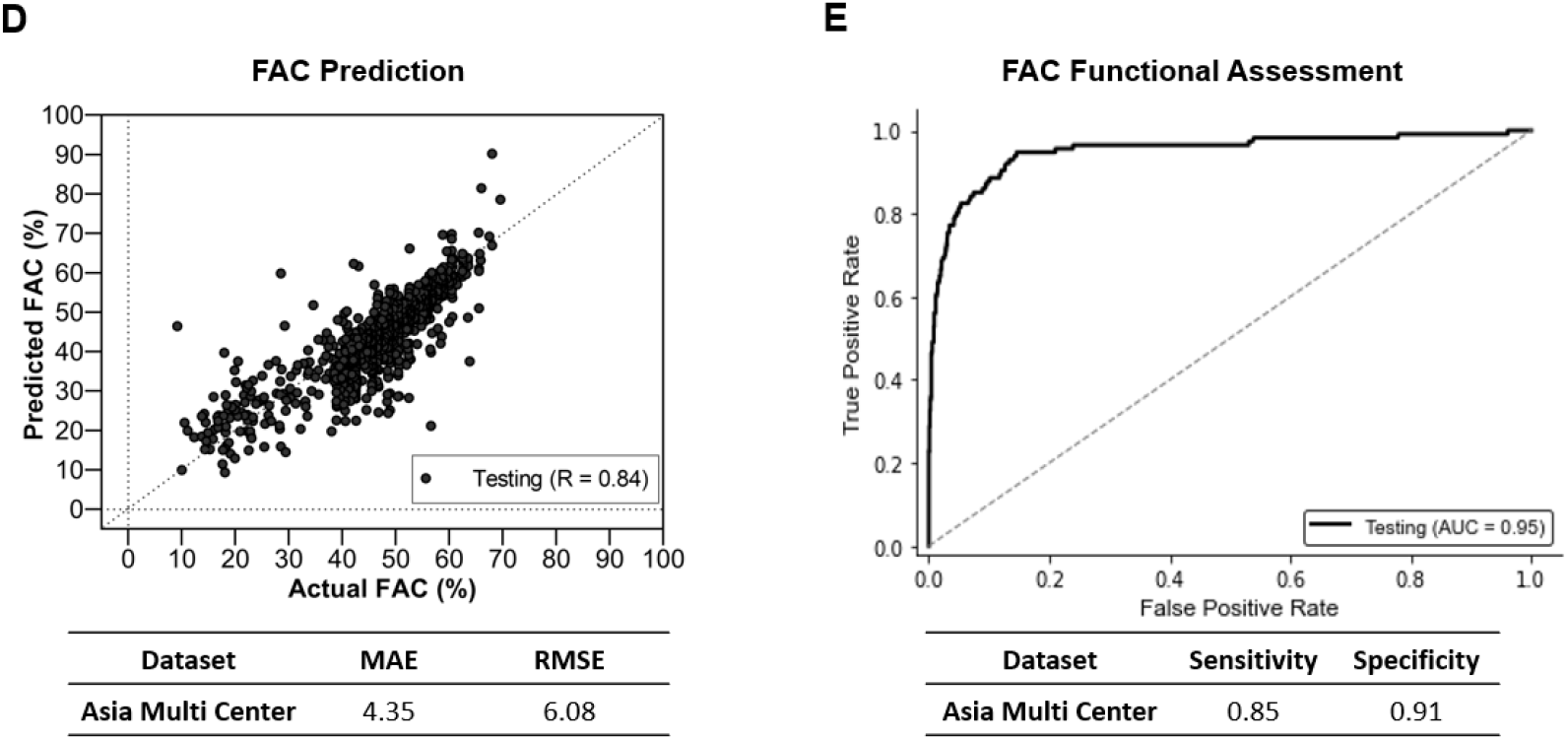
RV function assessment powered by AI learning of multi-center pediatric echocardiograms. A. Performance analysis, on U.S. single-center data (training and validation sets), of the Dice similarity coefficient for the overall, end-systolic, and end-diastolic tracing. B. The predicted RV FAC compared with the reported FAC for the U.S. single-center data (training and validation sets). MAE: Mean Absolute Error. RMSE: Root Mean Square Error. C. Receiver-operating characteristic curves for the RV function assessment with reduced RV FAC (<35%) for the U.S. single-center data (training and validation sets). D. The predicted RV FAC compared with the reported FAC for the Asia multi-center validation data. E. Receiver-operating characteristic curves for the RV function assessment with reduced RV FAC (<35%) for the Asia multi-center validation data.

A notable difference between the cohorts lies in the ratio of abnormal to normal patient videos. In the U.S. cohort, this ratio was 1.87, compared to 0.31 in the Asian cohort. This substantial disparity in data distribution likely contributed to the strong correlation observed in the Asian cohort, as the higher proportion of abnormal cases may have enhanced the model’s ability to distinguish between normal and abnormal RV function.

### RV-based AI learning to allow automated PH identification

Figure 5 demonstrates the RV-based classification model performance for CSN#1 and CSN#2 across the U.S. (A) and Asian (B) cohorts. In the U.S. cohort (A), CSN#1 achieved 95% accuracy in classifying normal cases and 73% for PH cases, with low misclassification rates (4.5% for normal and 18% for PH). CSN#2 demonstrated superior performance, correctly classifying 100% of cases with no misclassification or undetermined results. These findings highlight the model’s potential for broader applications in pediatric cardiology. In the Asian cohort (B), the left panel shows CSN#1’s performance in normal vs. PH classification, achieving 90% accuracy for normal cases and 86% for PH cases, with misclassification rates of 10% for normal and 24.9% for PH. The right panel shows CSN#2’s results for PH vs. TOF classification, where performance was with 54.5% of PH cases correctly classified and 33% of TOF cases undetermined.

**Figure 5.**
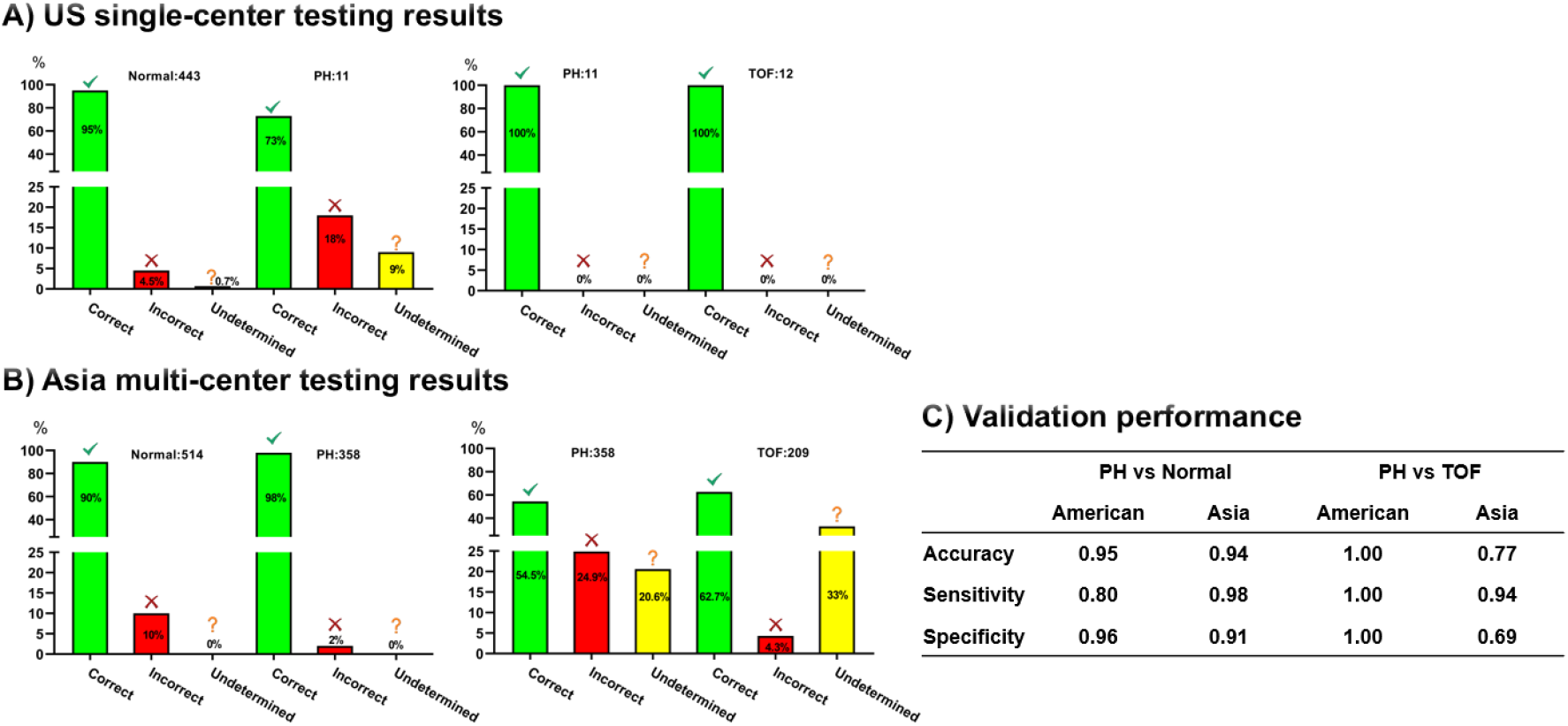
RV-based classification model performance: CSN#1 modeling detection of PH from normal conditions and CSN#2 modeling differentiating PH from TOF. A. U.S. single-center testing results; B. Asia multi-center testing results; C. validation performance measures.

Overall, CSN#1 demonstrated strong performance in distinguishing normal from PH, particularly in the U.S. cohort, while CSN#2 showed greater challenges in PH vs. TOF classification, especially in the Asian cohort. These differences highlight the variability in model performance across cohorts and tasks.

### Combining A4C and PSAX echocardiograms for RV learning based PH identification

The performance of automated echocardiogram segmentation for pediatric right ventricle assessment using US single center PSAX datasets is presented in Figure S7. Dice coefficients, a metric for segmentation accuracy, were calculated for three categories: Overall, End systole, and End diastole. The results demonstrate consistently high segmentation accuracy across all categories. Overall performance achieved Dice coefficients of 0.90 and 0.88, while End systole segmentation showed similar accuracy with coefficients of 0.90 and 0.87. Notably, the End diastole segmentation exhibited the highest accuracy, with Dice coefficients of 0.91 and 0.89. These results, ranging from 0.87 to 0.91, indicate robust performance of the automated segmentation method in delineating right ventricular structures in pediatric echocardiograms, with particularly strong performance during end diastole.

The comparison of A4C, PSAX, and ensemble models for predicting PH, normal cases, and TOF highlights the ensemble model’s superior performance (Figure S8). In distinguishing PH from normal cases, the ensemble model achieved robust accuracy (99%) and specificity (100%), while maintaining a sensitivity of 83%, similar to the A4C model. When differentiating PH from TOF, the ensemble model demonstrated robust accuracy (92%) and specificity (100%), outperforming PSAX in specificity. Although PSAX had the highest sensitivity (100%) for PH vs. TOF, its lower specificity (71%) indicates more false-positive results. The A4C model consistently balanced sensitivity and specificity but was outperformed by the ensemble in overall accuracy and specificity. These results confirm that integrating A4C and PSAX predictions into an ensemble framework improves diagnostic precision.

### Exploration of RV AI learning framework for LV Function assessment

Figures S9 and S10 illustrate model performance for LV EF prediction. In the U.S. cohort (Figure S9, single-center data), the model achieved a DICE coefficient of 0.91 for segmentation accuracy, with AUC values of 0.98 (training) and 0.92 (validation) in functional assessment. For direct comparison with Reddy et al.’s EchoNet-Peds^8^, we evaluated our U^2^Net based method using the Asia multi-center cohort (Table S3). Our approach demonstrated improved accuracy, evidenced by significantly lower MAE and RMSE values. A box plot highlights the predicted EF deviations from true values, showing our method’s tighter distribution around zero, reflecting greater precision, while EchoNet-Peds exhibited wider prediction deviations. The comparison’s statistical significance is confirmed by a p-value < 10^−8^, underscoring our method’s enhanced LV EF prediction capability. Figure S11 summarizes additional performance benchmark analysis of models in the U.S. single-center cohort.

## Discussion

This study introduces a video-based deep learning algorithm for comprehensive pediatric cardiac assessment, excelling in RV FAC prediction and FAC-based functional assessment. The model also performs advanced RV-based PH identification, highlighting its clinical relevance. Additionally, the algorithm extends to LV EF prediction by incorporating child growth-related parameters, enhancing LV functional assessment. Validation across ethnically diverse, multi-center pediatric cohorts in the United States and Asia revealed negligible racial effects, confirming its wide applicability. These findings establish the multimodal AI framework as a promising solution to key challenges in pediatric ventricular function assessment. Using a single Nvidia A100 GPU, the framework completes tasks in real time, with predictions taking only 0.013 seconds per frame, significantly faster than human assessments.

### Interpretation of Results

Our algorithm introduces a novel workflow for segmenting the RV, predicting FAC, and assessing RV function. Unlike traditional human assessments relying on single representative beats, our model evaluates cumulative beat-to-beat data, capturing variability, reducing missed abnormalities, and enhancing diagnostic precision. While focused on RV function, the algorithm also improves LV function assessment, broadening clinical utility. By automating these processes, it enhances accuracy, consistency, and reproducibility while addressing intra-observer variability.

Classification models differentiate normal, PH, and TOF cases using beat-by-beat RV segmentation and echocardiogram video analysis. The model’s ability to detect RV geometry changes associated with PH highlights its diagnostic utility. Distinguishing PH from TOF, though less clinically relevant, validates the algorithm’s robustness in identifying subtle RV morphological and functional differences, ensuring effectiveness in more complex conditions.

### Contextualization

Our findings extend prior research in pediatric echocardiography and AI cardiology applications. Assessing RV function is challenging due to its complex geometry and interpretive variability. The beat-to-beat analysis performed by our model overcomes these limitations, providing detailed and consistent evaluations. The additional LV function assessment capability enhances the model’s clinical relevance, supporting holistic cardiac evaluations.

Cross-center validation across U.S. and Asian institutions underscores the model’s generalizability in diverse clinical settings without fine-tuning. This scalability addresses global variability in healthcare resources and patient demographics, positioning the model as a transformative tool for pediatric cardiac care.

### Strengths and Limitations

The model’s strengths include automating labor-intensive pediatric RV and LV tracing, reducing manual workload, and adhering to clinical guidelines. Comprehensive beat-to-beat analysis ensures detailed and reliable assessments, surpassing traditional single-beat evaluations. These attributes enhance diagnostic accuracy, reduce observer variability, and streamline clinical workflows, improving pediatric care.

Limitations include the algorithm’s training on high-quality echocardiographic videos from a U.S. academic center, which may not represent real-world variability. Preliminary results suggest robustness to different video qualities, but broader validation is needed, particularly in low-resource settings. Additionally, while LV assessments were included, they were secondary to RV analysis and warrant further investigation. Addressing these limitations will be critical for ensuring reliability and applicability across diverse healthcare contexts.

### Clinical Relevance and Implications

Our model’s clinical implications are significant, particularly in automating pediatric RV function assessment. Real-time predictions facilitate earlier detection of subclinical RV changes, enabling timely interventions and improved management of pediatric cardiac conditions. Automating beat averaging, a guideline-recommended practice, enhances consistency and quality in RV and LV assessments, contributing to better clinical outcomes for children with cardiac conditions.

### Future Directions

Future research should prioritize validating the algorithm in real-world clinical settings with varying video quality, acquisition techniques, and operator expertise. Expanding validation to low-resource environments is essential for global applicability. Integrating echocardiogram AI learning with EMR systems has demonstrated potential, and incorporating multimodal datasets (e.g., echocardiograms, EMRs, clinical data) could further refine the framework. Extending multitask capabilities to include additional pediatric cardiac abnormalities and advanced features will enhance the model’s clinical impact.

## Conclusion

This study advances pediatric ventricular function assessment, focusing on RV function while offering additional LV insights. Leveraging video-based deep learning, our model improves precision in detecting cardiac dysfunction, enabling earlier intervention and better management of pediatric heart conditions. Its robust performance across diverse populations highlights its potential to transform pediatric cardiac care. Continued refinement and validation will ensure broader adoption, facilitating frequent and accurate cardiac evaluations and improving outcomes for children with heart diseases. The development of scalable, efficient AI-driven tools represents a pivotal step toward personalized pediatric healthcare.

## Data Availability

The data and code used in this work will be publicly available on GitHub (https://github.com/bxlinglaboratory/EchoRV) upon publication.

## Author contributions

CR and XBL conceptualized the study in collaboration with DBM;

CR, YY, MQ, YT, HX, SY, QW, LPW, LJF, ZYJ, YJY, YQZ, HZ contributed to patient enrollment and sample collection for the NCT trial;

BJ, ZH, YL, SZ, QT, EL, and XBL developed AI learning framework and statistical tools;

NO, TI, ZH, and XBL analyzed the data and performed model development;

CR, ZH, NO, TI, RJW, IM, NA, BG, MSA, KGS, HJC, GLD, DKS, AT, HC, SC, DBM, and XBL contributed to the writing and editing of the manuscript.

## Code and data availability

Data and code used in this work are available at https://github.com/bxlinglaboratory/EchoRV,

## Acknowledgments

BJ, YL, QT, and SZ are employees of HBI Solutions Inc. NO and TI are Stanford University visiting scholars from Nippon Life Insurance Company. However, the companies had no role in the design of the study, data collection, data analysis, interpretation, or the preparation of this manuscript. All research and experimental work were conducted independently, and no external influence impacted on the study’s findings or conclusions. AT reports consulting fees from Siemens Healthineers for work unrelated to echocardiography. The other authors associated with hospitals and universities declare no competing interest.

## Data sharing

All potentially identifiable data were encrypted to protect anonymity. Access to the data is restricted to investigators whose proposals are approved and who have signed a data access agreement.

## Notes

### Author Declarations

The study protocol was reviewed and approved by the Institutional Review Boards of all participating hospitals (Stanford Children's Hospital, the Children's Hospital of Chongqing Medical University, Chongqing YouYou BaoBei Women's and Children's Hospital, and Shanghai Children's Medical Center), ensuring strict adherence to institutional guidelines. To safeguard patient privacy and confidentiality, comprehensive data protection measures were implemented. These included the removal of personal identifiers, secure file format conversion, and thorough manual dataset review. These rigorous procedures ensured compliance with ethical standards for managing sensitive medical information.

