## Supplementary Materials for "AI Learning for Pediatric Right Ventricular Assessment: Development and Validation Across Multiple Centers"

**Supplementary Materials and Methods**

### Supplementary Materials and Methods

#### Study population

This study integrated data from two distinct cohorts.

- **U.S. Single-Center Cohort:**
  Data were collected from children aged 0 to 18 years enrolled at Stanford Children's Hospital between 2014 and 2020, with each participant contributing at least one cardiogram video. Within this cohort, the average number of echocardiograms per normal patient was 4.90, while for abnormal patients, the average was 51.4.
- **Asian Multi-Center Cohort:**
  Data were gathered between 2015 and 2024 from three institutions: the Children's Hospital of Chongqing Medical University, Chongqing YouYou BaoBei Women's and Children's Hospital, and Shanghai Children's Medical Center. In this cohort, the average number of echocardiograms per normal patient was 2.07, compared to 14.84 for abnormal patients.

The study protocol was reviewed and approved by the Institutional Review Boards of all participating hospitals, ensuring strict adherence to institutional guidelines. To safeguard patient privacy and confidentiality, comprehensive data protection measures were implemented. These included the removal of personal identifiers, secure file format conversion, and thorough manual dataset review. These rigorous procedures ensured compliance with ethical standards for managing sensitive medical information.

Two groups of children are included. Group I: Children with normal right ventricular anatomy and function assessment based on screening echocardiograms and physician follow up. Group II: Children with right ventricular abnormalities who have potential risk of adverse outcomes. The two groups are age-, gender-, and size-matched. The inclusion criteria for Group II are:

1. Patients aged 0-18 years old.
2. Patients with the following diagnoses (defined as abnormal RVs): premature infants with lung disease, congenital heart disease with systemic right ventricles, surgically repaired congenital heart disease resulting in pressure and/or volume load on the RV (tetralogy of Fallot, Double outlet right ventricle, etc.), and idiopathic pulmonary hypertension.

Each echocardiogram consists of multiple video clips of various parts of the heart. We will select a fraction of the clips, focusing on the apical-four-chamber (A4C), focused RV views, short axis RV views, and RV inflow-outflow views. A4C views were selected as the primary focus due to their relevance in RV assessment. Given the unique challenges of RV imaging—complex geometry, positioning, and absence of standardized landmarks, data were collected by sonographers with specialized training in cardiac imaging, following established protocols to ensure image quality and consistency and the data were stored securely with metadata, including view types and measurements. Our cardiologists would label the normal and abnormal studies with commonly accepted right ventricular measurements as per American Society of Echocardiography guidelines. The image quality of all the studies will also be classified as: 1) good; 2) average; 3) poor; or 4) not analyzable. Each potential frame from which to measure RV characteristics would also be labelled as 1) optimal; and 2) sub-optimal.

Uniform image processing techniques were employed to standardize the collected videos, thereby mitigating variations in quality across different centers. The sampling dates and relevant clinical information were collected as part of the data collection process.

#### U^2^-Net based RV segmentation

The model design and training were implemented in Python 3.10.9 using the PyTorch 2.3.1 deep learning library, which offers a robust framework for building and training neural networks. To achieve automated segmentation of RV regions, a U^2-^Net-based deep learning framework^1^ was employed.

This model architecture includes five encoders and four decoders, allowing it to capture intricate spatial details. Training was performed using pixel-level binary cross-entropy loss, optimizing the segmentation performance. We initialized the model with random weights and fine-tuned it using the adaptive moment estimation (Adam) optimizer. The initial learning rate was set to 1e-4 and adjusted during training to ensure effective convergence. Pretrained weights from U^2^-Net were also applied to further refine our model, providing a strong starting point for training. Additionally, we repeated our previous segmentation architecture with EchoNet-Peds (Deeplabv3+^2^) to compare the two deep learning performances.

The framework operates on frame-level echocardiographic video data, extracting systolic and diastolic frames for segmentation. The U^2^-Net model accurately delineates the ventricular contours, enabling precise segmentation of the RV in both phases of the cardiac cycle. Following segmentation, beat-by-beat analyses were performed by tracking temporal changes in the segmented ventricular areas across frames. This approach provides critical measurements of ventricular function, such as systolic and diastolic areas (s1, s2, etc., and d1, d2, etc.), which form the basis for quantitative assessments of cardiac performance. The method's robustness ensures reliable segmentation across diverse echocardiographic views and clinical conditions.

#### RV FAC prediction and function assessment

This study utilized the RV FAC values calculated by on-site cardiologists, as documented in the finalized clinical reports, as the gold standard (ground truth) for training and benchmarking our AI learning algorithms. Using segmentation models, FAC was calculated from area changes across frames. Peaks and valleys in area values, representing systolic and diastolic frames respectively, were identified via Savitzky-Golay filtering for noise reduction. FAC was calculated as follows:

$$\left\{ \begin{aligned} {FAC}_{n}=\frac{d_{n}-s_{n}}{d_{n}},num\left( d \right)\geq num(s) \\ {FAC}_{n}=\frac{d_{n}-s_{n-1}}{d_{n}},num\left( d \right)\geq num(s) \end{aligned} \right.$$

Where $\boldsymbol{s}_{\boldsymbol{n}}$ represents the area value of the $n^{th}$ systolic frame and $\boldsymbol{d}_{\boldsymbol{n}}$ represents the area value of the $n^{th}$ diastolic frame. The variable "n" denotes the $n^{th}$ beat cycle of the video, and "num()" represents the number of diastolic or systolic frames in the video.

Table S2 summarizes the characteristics and distribution of RV FAC prediction data across training and testing cohorts. The U.S. cohort was split into training (624 patients) and testing (240 patients) subsets, while the Asia cohort included 835 patients in the testing set. Median age was similar between the training (9.3 years, IQR 5–14) and testing (9.1 years, IQR 4–13) groups, but lower in the Asia cohort (3.6 years, IQR 0.8–4.6). The mean FAC was 46.5% (IQR 41.8–50.8) for U.S. training, 47.5% (IQR 42.6–52.1) for U.S. testing, and 46.9% (IQR 42.5–52.5) for Asia testing. Diastolic and systolic areas were consistent in the cohorts, highlighting the cohort's multi-center, multivariable nature and supporting the model’s robustness in learning transfer.

Figure 2 illustrates our RV AI workflow to predict FAC and allow automated assessment of RV function using single-center and multi-center echocardiograms. The U.S. single-center cohort was split into a training dataset and validation dataset (Step 1). A deep learning model, U^2^-Net, was then trained using the U.S. single-center training data to perform automatic RV frame-level beat-by-beat segmentation (Figure S2) on echocardiographic videos (Step 2). The segmentation outputs were used to calculate RV FAC (Step 3), a key measure of RV function derived from the area change between end-diastole and end-systole across multiple video frames. The extracted FAC values were then used to predict the probability of abnormal RV function (Step 4). The model’s performance was evaluated based on the accuracy of these predictions. To ensure generalizability across populations, the trained model was validated using test datasets from both the U.S. single-center and Asia multi-center cohorts (Step 5). The diagnostic performance was quantified using the Area Under the Receiver Operating Characteristic (ROC) curve, measuring the model’s ability to distinguish between normal and abnormal RV functions based on FAC predictions (Step 6).

#### FAC prediction performance by gender

The scatter plots in Figure S12 illustrate the predicted versus actual FAC (Fractional Area Change) values stratified by gender, with panel (A) representing males and panel (B) representing females. The results demonstrate strong predictive performance, with testing correlation coefficients of R = 0.83 for males and R = 0.90 for females. Both plots exhibit a close alignment of data points along the diagonal line, indicating accurate predictions for both genders. The findings suggest that the model is robust in predicting FAC across gender groups, with slightly higher accuracy observed in females.

#### RV based CSN modeling for pulmonary hypertension (PH) identification

Building on the above RV segmentation, we further developed deep-learning classification models to identify PH using multi-center pediatric RV echocardiograms.

We employed the Channel Separated Convolutions Network (CSNs)^3^ and its 3D variant for automated PH identification based on RV echocardiogram analysis. CSNs are inspired by the Xception architecture^4^, which factorizes 2D convolutions into channel and spatial dimensions for object classification. CSNs take this further by factorizing 3D convolutions into channel and space-time dimensions, making them ideal for tasks like action recognition. These networks use bottleneck blocks and group/depthwise convolution to enhance efficiency.

The classification models were trained using a stochastic gradient descent optimizer, with an initial learning rate of 1e-4, a momentum of 0.9, and a batch size of 32. Training was conducted over 5 epochs, and the weights from the epoch with the lowest validation loss were chosen for final testing.

For the model input, video clips of 64 frames were used, starting from the 0^th^ frame, with a frame sampling period of 2, determined through hyperparameter tuning. Selecting the optimal clip length and sampling period is critical for capturing relevant temporal information from the echocardiogram videos.

The workflow (Figure 3) begins with the construction of a training cohort from a U.S. single center (Step 1), the training data is used to train a U^2^-Net model for RV segmentation in cardiac imaging (Step 2). The segmented images undergo beat-by-beat analysis (Step 3), extracting relevant spatiotemporal data (Step 4). This data is processed by two channel separated convolutional neural networks (CSN #1 and CSN #2) for classification: CSN #1 differentiates between normal and PH cases, while CSN #2 differentiates between PH and TOF cases. The remaining U.S. single-center data is reserved for validation. The trained models are then tested on both the U.S. single-center data and a multi-center cohort from Asia (Step 5). Finally, the performance of the models is evaluated on the multi-center validation cohort (Step 6). Solid arrows represent the training workflow, while dashed arrows indicate the validation process.

Table S3 outlines patient and video characteristics for the PH identification learning. The dataset included three sets: Single Center Training, the U.S. single center testing, and Asia multi-center testing. In total, the U.S. single center training set includes 1,232 patients, with an interquartile range (IQR) of age spanning from 5 to 14 years, while the U.S. Single center testing set has 466 patients with a similar age range (IQR: 5–14 years). The Asia multi-center testing set, with 1,098 patients, has a younger patient cohort with an age IQR of 1.5 to 7.0 years. Patient demographics and video distributions showed notable variability, especially in the proportion of TOF cases in the Asian dataset (9,763 TOF videos), underscoring the diversity of the dataset.

While the A4C view is a cornerstone^5^ for assessing RV size, shape, and systolic function, it does not capture the infundibular region. To address this limitation, the parasternal-short-axis (PSAX) view is often used alongside the A4C view to provide a more comprehensive evaluation. In this study, we investigated the combined analysis of A4C and PSAX views (Table S4, Figures S3, S4) to assess RV function and enable automated identification of pulmonary hypertension (PH).

#### Comparison of learning methods

Figure S11 presents a benchmark analysis of models in the U.S. single-center cohort. Panels A and B highlight the superior performance of the our U²-Net implementation to allow robust LV and RV segmentation during training and validation, achieving the lowest loss values compared to U²-Net and DeepLab v3+. Panel C shows the RV PH/TOF classification model's performance with video inputs of 32, 48, and 64 frames, where the 64-frame model demonstrates the most consistent and significant loss reduction. The table outlines model architecture, with the modified U²-Net featuring fewer encoder and decoder blocks (RSU: 4 and 3) for a streamlined yet high-performing design. All experiments used binary cross-entropy (BCE) as the loss function.

### Supplementary Figures

#### Figure S1. Venn diagram of multi-center study cohorts.

(A) Venn diagram illustrating the distribution of **patient counts** across four studies. Overlapping regions indicate patients shared among multiple studies, while non-overlapping sections represent unique patients within each study. (B) Venn diagram depicting the **echocardiogram video counts** per study, highlighting shared and unique video contributions from each study site. The distribution demonstrates the overlap in echocardiographic data availability across the multi-center cohorts.

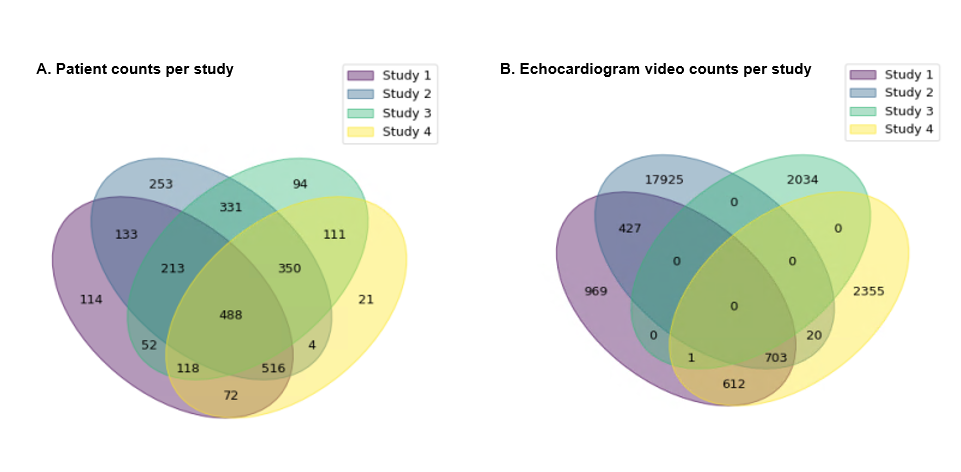

#### Figure S2. RV frame-level segmentation and beat-by-beat AI learning.

(A) **RV Frame-Level Segmentation:** Illustration of the neural network-based right ventricular (RV) segmentation process applied to echocardiography videos. The model extracts systolic and diastolic frames to delineate RV boundaries, enabling precise assessment of RV function. (B) **RV Beat-by-Beat Analysis:** A continuous segmentation area curve across frames, demonstrating the beat-by-beat variations in RV contraction and relaxation. Peaks (d1, d2, d3) correspond to diastole, and troughs (s1, s2, s3, s4) correspond to systole, capturing the dynamic changes in RV function over multiple cardiac cycles.

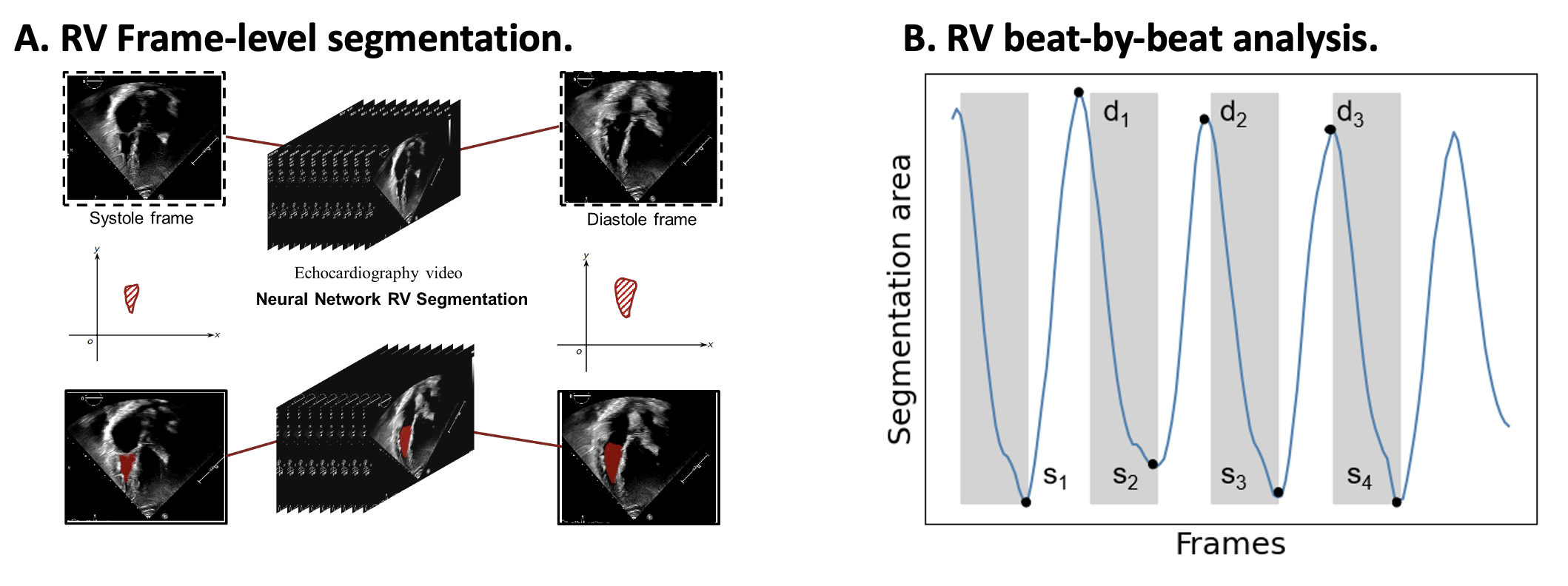

#### Figure S3. Combining A4C and PSAX view for AI learning to allow RV Function assessment and PH identification.

This schematic illustrates the AI-based right ventricular (RV) function assessment and pulmonary hypertension (PH) identification using both apical four-chamber (A4C) and parasternal short-axis (PSAX) echocardiographic views. Solid lines represent training workflows, while dashed lines indicate validation workflows.

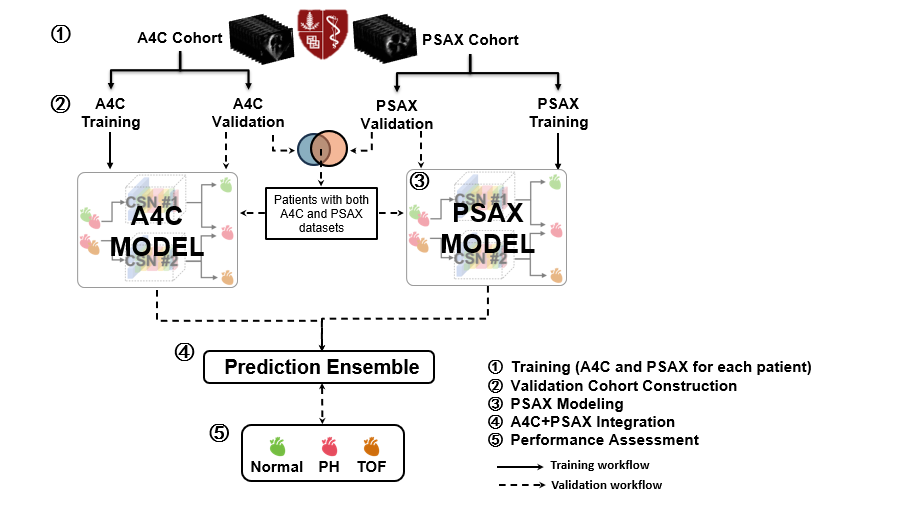

#### Figure S4. Demonstration of segmentation output for U^2^-net

This figure presents segmentation results using the U²-Net model for different cardiac conditions—Normal, Pulmonary Hypertension (PH), and Tetralogy of Fallot (TOF)—across two echocardiographic views: Apical 4 Chamber (A4C) and Parasternal Short Axis (PSAX). Raw Image Row displays original echocardiographic frames for each condition and view; Ground Truth Row shows manually labeled segmentation masks as reference; Segmentation Row displays the U²-Net model's predicted segmentation output.

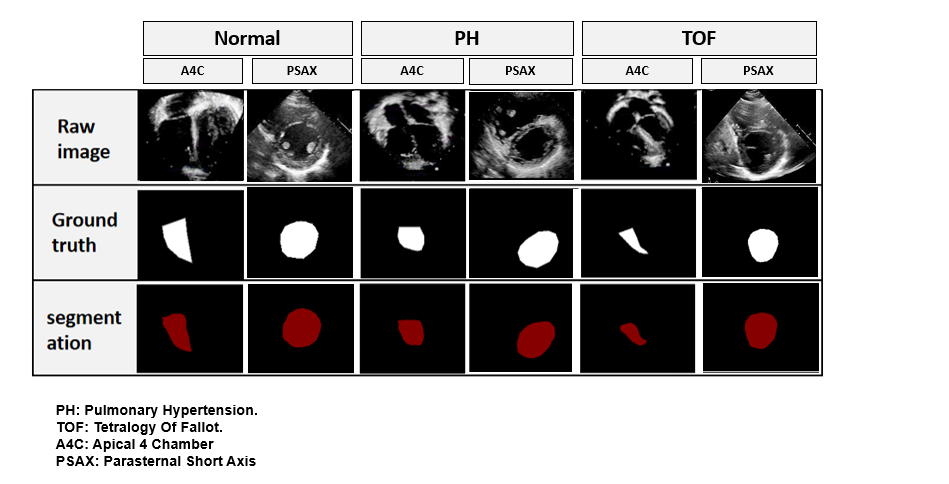

#### Figure S5. Exploration of the RV AI learning framework for LV segmentation with beat-by-beat analysis of segmentation areas.

**(A)** LV Frame-level segmentation: Example frames from systole and diastole showing LV segmentation using a neural network-based model. The segmented LV areas are highlighted in red. **(B)** LV Area change curve: The cyclic variation in the LV segmentation area is shown over time, with systolic (s₁, s₂, s₃) and diastolic (d₁, d₂, d₃) phases indicated.

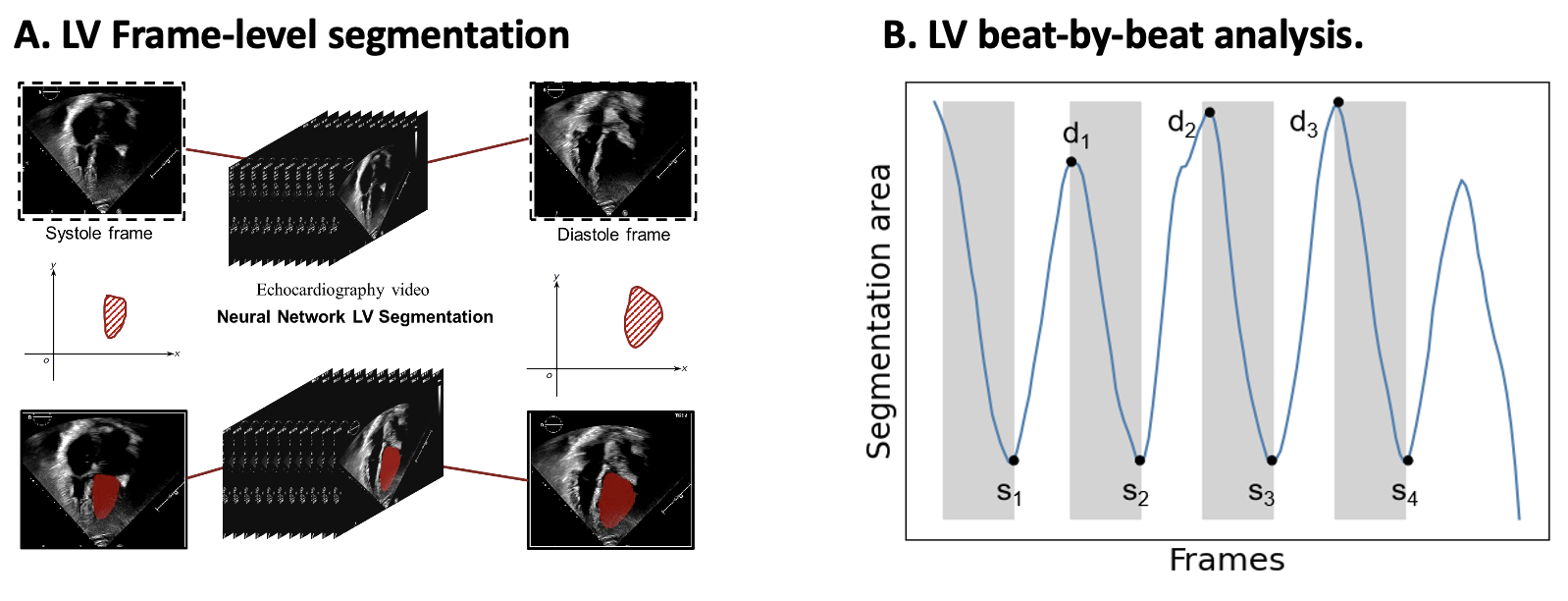

#### Figure S6. The automated LV function assessment pipeline on pediatric echocardiograms.

Automated left ventricle function assessment from US single-center and Asia multi-center pediatric echocardiograms. EF: ejection fraction. LAC: LV 2D Area Change

The workflow begins with cohort construction (①), where data from a U.S. single-center and Asia multi-center pediatric cohorts are used for training and validation. The U2-Net model (②) is trained to perform LV segmentation, followed by LAC (LV 2D Area Change) extraction from the segmented echocardiograms (③). The extracted LAC, along with patient factors such as age, sex, and heart rate, is used to estimate the EF (④). The model is validated on multi-center cohorts (⑤) and the performance is assessed by calculating the area under the curve (⑥), evaluating the model's accuracy in detecting abnormal EF values.

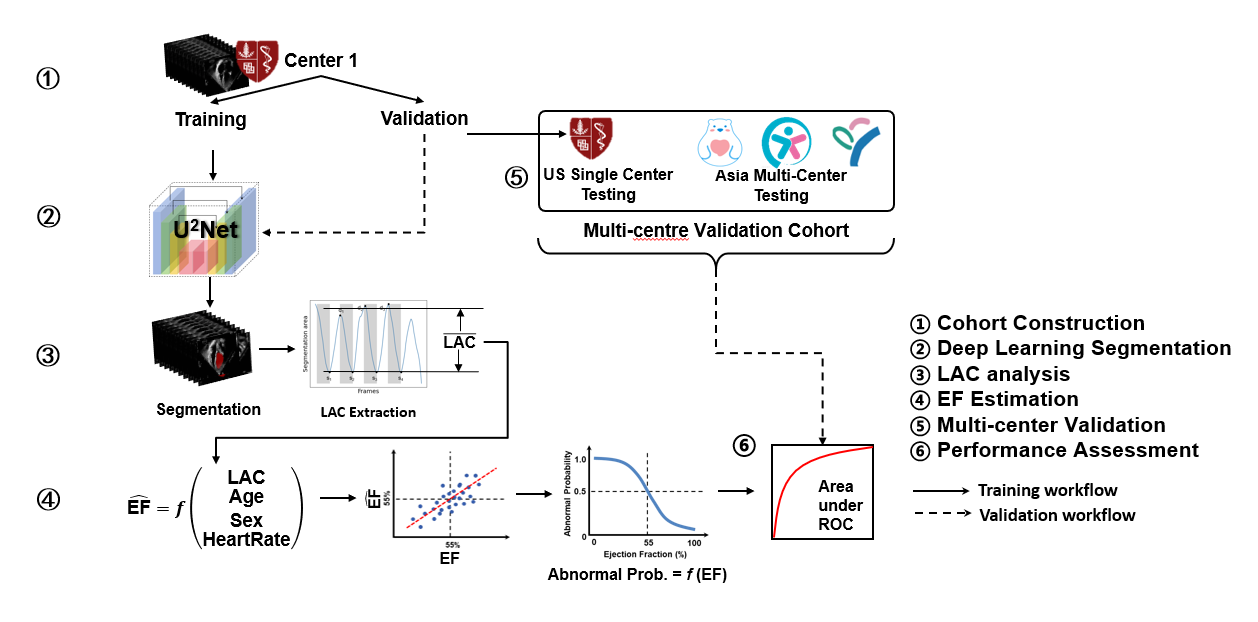

#### Figure S7. Performance of automated echocardiogram segmentation for pediatric right ventricle assessment with US single center PSAX datasets.

The bar chart illustrates the DICE coefficient performance of an automated echocardiogram segmentation model for pediatric right ventricle (RV) assessment using a single-center PSAX dataset. The DICE coefficient measures segmentation accuracy, with higher values indicating better agreement with ground truth. The comparison is made across three categories: Overall, End Systole, and End Diastole. The blue bars represent one segmentation approach, while the red bars represent another for comparison.

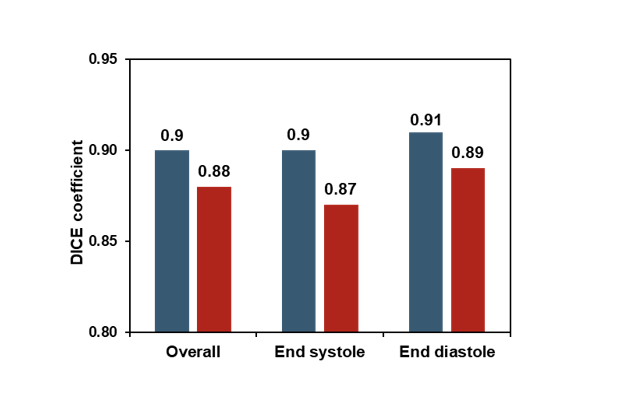

#### Figure S8. A4C, PSAX and A4C/PSAX based model performance comparison.

(A-C) Bar plots showing classification performance across different echocardiographic views: (A) Apical Four-Chamber (A4C), (B) Parasternal Short-Axis (PSAX), and (C) Ensemble model combining both views. Each subplot presents classification results for Normal vs. Pulmonary Hypertension (PH) and PH vs. Tetralogy of Fallot (TOF) cases. Green bars represent correctly classified cases, while red bars indicate misclassified cases. Percentages indicate classification accuracy for each group. (D) Summary table of validation performance metrics, including accuracy, sensitivity, and specificity, for distinguishing PH from Normal and PH from TOF across A4C, PSAX, and Ensemble models. The Ensemble approach demonstrates the highest overall accuracy in distinguishing PH from Normal (0.99) and improved sensitivity in PH vs. TOF classification.

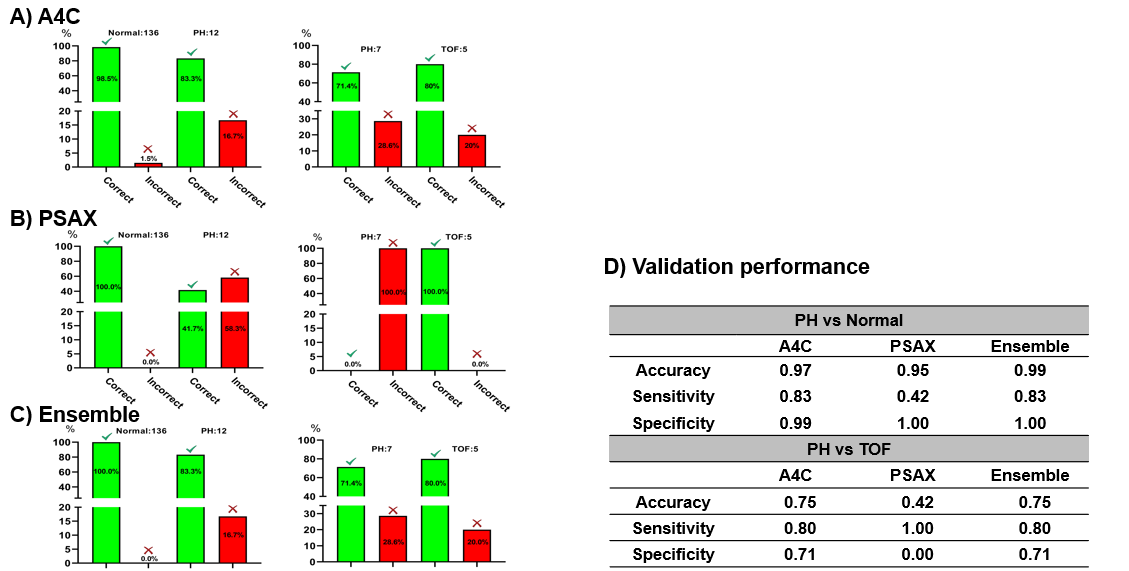

#### Figure S9. LV function assessment powered by RV based AI learning of multi-center pediatric echocardiograms.

A. Performance analysis, on U.S. single-center data (training and validation sets), of the Dice similarity coefficient for the overall, end-systolic, and end-diastolic tracing. B. The predicted LV EF compared with the reported EF for the U.S. single-center data (training and validation sets). MAE: Mean Absolute Error. RMSE: Root Mean Square Error. C. Receiver-operating characteristic curves for the EF function assessment with reduced LV EF (<45%) for the U.S. single-center data (training and validation sets).

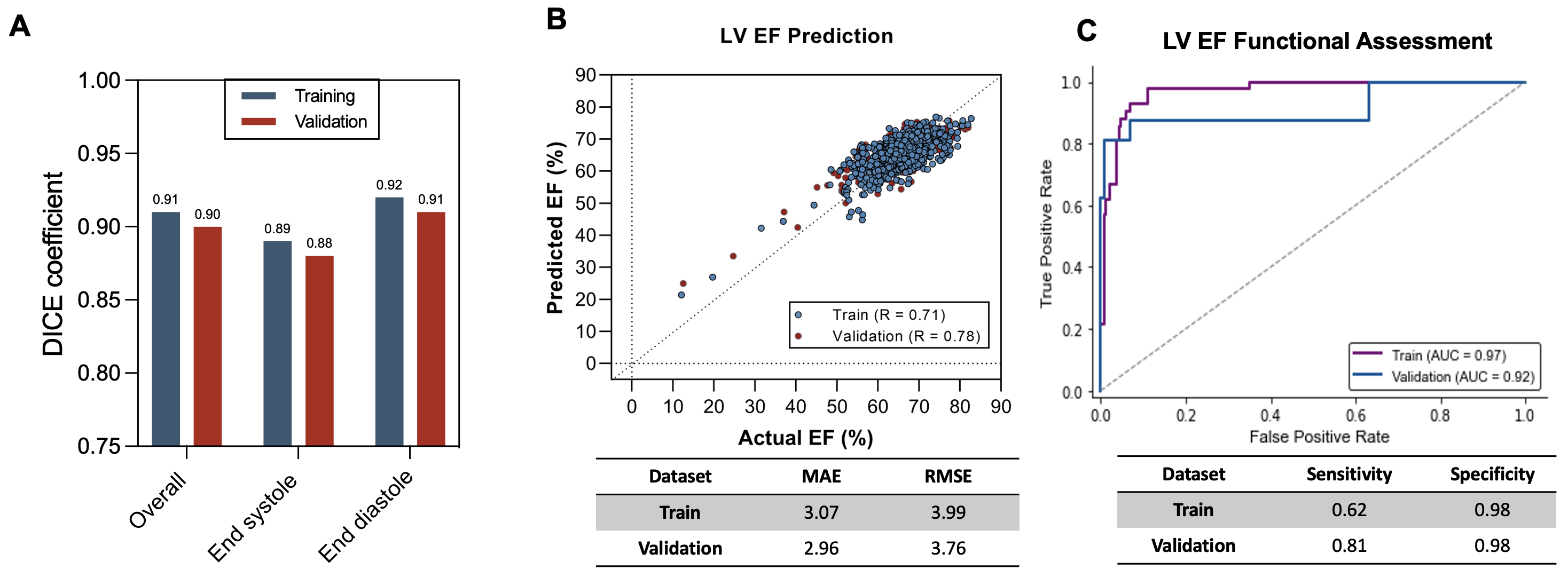

#### Figure S10. Asia multi-center LV EF prediction comparing algorithms of EchoNet-Peds and our RV-based learning framework.

The figure compares the performance of EchoNet-Peds and U²-Net in predicting left ventricular ejection fraction (LV EF) across a multi-center dataset from Asia. Left Panel (Bar Chart) displays Mean Absolute Error (MAE) and Root Mean Square Error (RMSE) for both models. Right Panel (Box Plot) shows the distribution of errors (Predicted EF - Ground Truth EF) for both models.

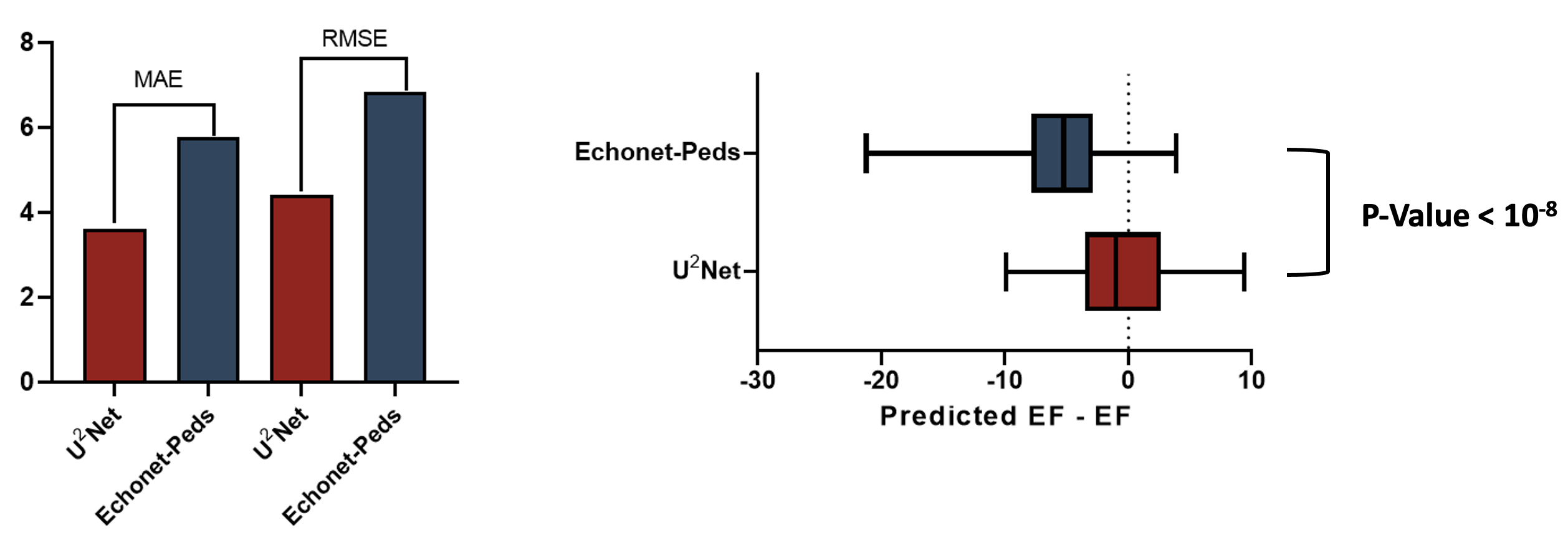

#### Figure S11. Benchmark analysis of different models in the U.S. single-center cohort

(A-B) Training and validation loss curves for (A) left ventricular (LV) and (B) right ventricular (RV) segmentation models. The performance of DeepLab v3+, U²-Net, and the modified U²-Net (this study) is compared. The modified U²-Net demonstrates the lowest loss, indicating superior segmentation performance. (C) Pretraining loss curves for the RV PH/TOF classification model, using input videos with 32, 48, and 64 frames. The model trained with 64-frame videos shows the most stable and lowest loss, suggesting improved learning efficiency. (D) Architectural comparison of different segmentation models, detailing the encoder and decoder configurations. The DeepLab v3+ uses a DCNN-based encoder, while the U²-Net and modified U²-Net (this study) utilize residual shuffle units (RSUs) with varying block depths. All models were trained using Binary Cross-Entropy (BCE) as the loss function

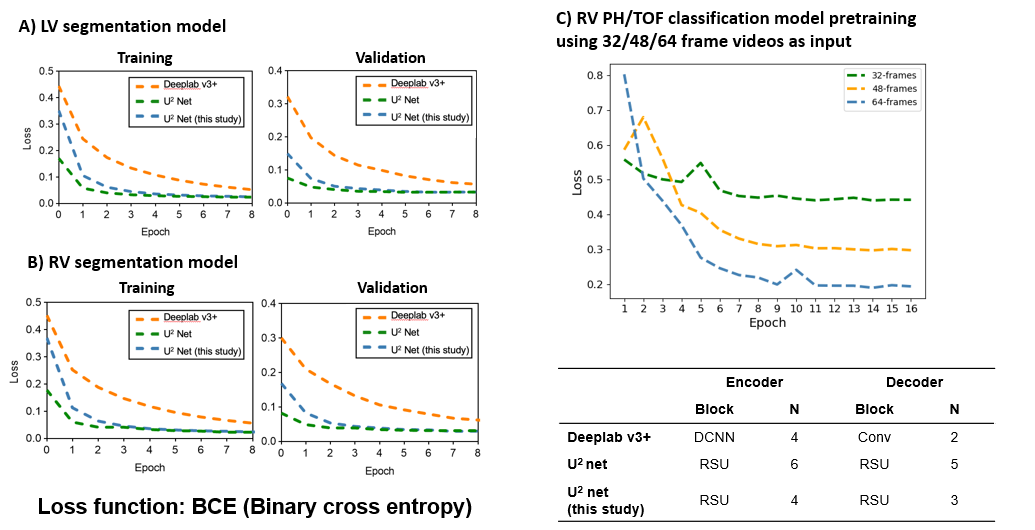

#### Figure S12. FAC Prediction Performance by Gender

Scatter plots comparing predicted fractional area change (FAC) with actual FAC values for (A) male and (B) female subjects. Each point represents an individual test sample. The dotted diagonal line indicates the line of perfect agreement (ideal prediction). The correlation coefficient (R) for testing data is 0.83 for males and 0.90 for females, indicating strong model performance in both groups, with slightly higher accuracy for females.

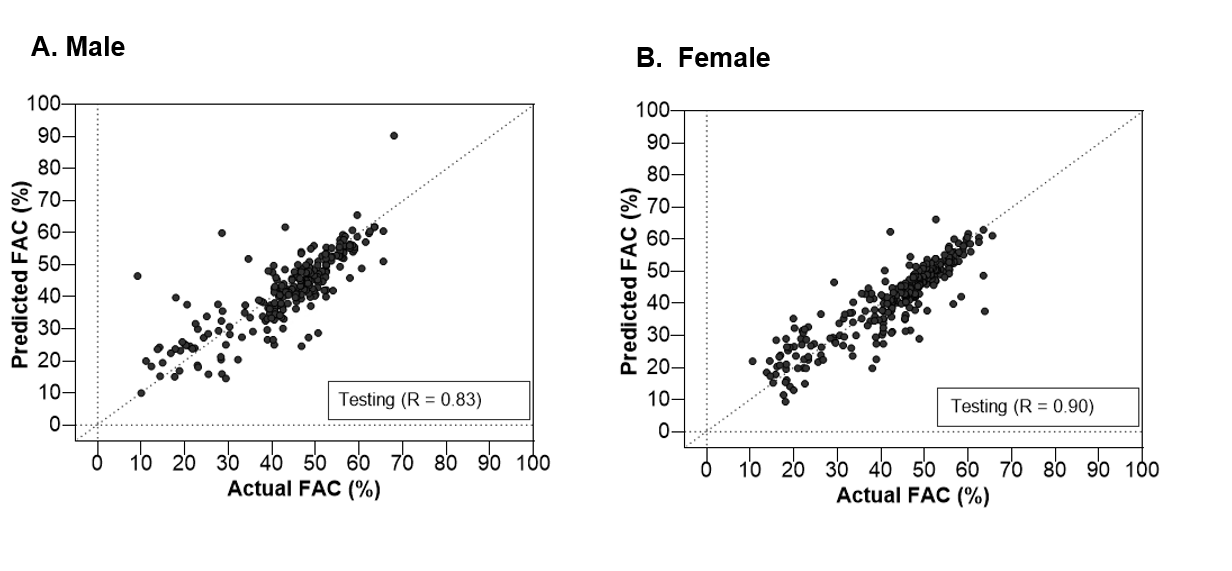

### Supplementary Tables

#### Table S1. Multicenter pediatric patient characteristics.

|  | **US Single Center** | **Asia multi-Center** |
| --- | --- | --- |
| **# Patients** | **1763** | **2230** |
| **Age (IQR) [year]** | **9.6 (5,14)** | **3.7 (0.9,4.7)** |
| **Gender** |  |  |
| **# Male** | **684** | **400** |
| **#  Female** | **492** | **480** |
| **# N.A.** | **587** | **1350** |
| **Patients Group** |  |  |
| **# Normal** | **1681** | **1531** |
| **# Pulmonary Hypertension (PH)** | **41** | **418** |
| **# Tetralogy of Fallot (TOF)** | **41** | **281** |
| **# All Collected Videos** | **13369** | **11615** |
| **# Normal** | **8862** | **1412** |
| **# Pulmonary Hypertension (PH)** | **1951** | **428** |
| **# Tetralogy of Fallot (TOF)** | **2556** | **9775** |

#### Table S2. Patient characteristics and videos for RV FAC prediction and function assessment

|  | **U.S. Single Center** | **U.S. Single Center** | **Asia Mult-center** |
| --- | --- | --- | --- |
|  | **Training** | **Testing** | **Testing** |
| **# Patients** | **624** | **240** | **835** |
| **All Collected Videos** | **968** | **399** | **1338** |
| **Age (IQR) [year]** | **9.3 (5,14)** | **9.1 (4,13)** | **3.6 (0.8,4.6)** |
| **Gender** |  |  |  |
| **# Male** | **356** | **124** | **196** |
| **#  Female** | **236** | **107** | **215** |
| **# N.A.** | **32** | **9** | **424** |
| **A4C Videos with RV FAC label** |  |  |  |
| **# Normal** | **856** | **373** | **1159** |
| **# Pulmonary**  **Hypertension (PH)** | **50** | **10** | **120** |
| **# Tetralogy of Fallot**  **(TOF)** | **62** | **16** | **59** |
| **FAC (IQR) [%]** | **46.5 (41.8,50.8)** | **47.5 (42.6,52.1)** | **46.9 (42.5,52.5)** |
| **Diastolic area (S.D.)**  **[cm^2^]** | **65.7 (10.2)** | **66.7 (24.0)** | **____** |
| **Systolic area (S.D.)**  **[cm^2^]** | **106.1 (13.8)** | **106.9 (13.2)** | **____** |

**S.D.: Standard deviation. IQR: the interquartile range.**

#### Table S3. Patient characteristics and videos for the identification of pulmonary hypertension (PH).

|  | **US Single Center** | **US Single Center** | **Asia Mult-center** |
| --- | --- | --- | --- |
|  | **Training** | **Testing** | **Testing** |
| **# Patients** | **1232** | **466** | **1098** |
| **All Collected Videos** | **5234** | **2699** | **11160** |
| **Age (IQR) [year]** | **9.5 (5,14)** | **9.6 (5,14)** | **4.7 (1.5,7.0)** |
| **Gender** |  |  |  |
| **# Male** | **519** | **203** | **127** |
| **#  Female** | **378** | **160** | **166** |
| **# N.A.** | **335** | **103** | **805** |
| **A4C Videos** |  |  |  |
| **# Normal** | **2544** | **1432** | **976** |
| **# Pulmonary**  **Hypertension (PH)** | **1200** | **495** | **421** |
| **# Tetralogy of Fallot**  **(TOF)** | **1490** | **772** | **9763** |

#### Table S4. U.S. single-center pediatric study with apical 4 chamber (A4C) and parasternal short axis (PSAX) view datasets.

|  | **PSAX Training** | | **A4C + PSAX Validation** |
| --- | --- | --- | --- |
|  | **Training** | **Validation** |  |
| **# Patients** | 1227 | 530 | 148 |
| **Normal** | 1167 | 508 | 139 |
| **PH** | 31 | 10 | 4 |
| **TOF** | 29 | 12 | 5 |
| **Age (IQR) [yr]** | 9.7 (5, 14) | 9.7 (5, 15) | 10 (5, 14) |
| **Gender** |  |  |  |
| **Male** | 680 | 301 | 86 |
| **Female** | 505 | 214 | 55 |
| **N.A.** | 42 | 15 | 7 |
| **# videos** | | |  |
| **Normal** | 1167 | 508 | A4C: 419  PSAX: 139 |
| **PH** | 120 | 34 | A4C: 15  PSAX: 10 |
| **TOF** | 149 | 56 | A4C: 39  PSAX: 29 |

#### Table S5. Patient characteristics and videos for LV EF prediction.

|  | **U.S. Single Center** | **U.S. Single Center** | **Asia Mult-center** |
| --- | --- | --- | --- |
|  | **Training** | **Testing** | **Testing** |
| **# Patients** | **756** | **311** | **612** |
| **All Collected Videos** | **1868** | **765** | **1068** |
| **Age (IQR) [year]** | **9.6 (5,14)** | **9.9 (6,14)** | **2.5 (0.5,3.0)** |
| **Gender** |  |  |  |
| **# Male** | **439** | **176** | **84** |
| **#  Female** | **309** | **133** | **109** |
| **# N.A.** | **8** | **2** | **419** |
| **A4C Videos with LV EF label** |  |  |  |
| **# Normal** | **1820** | **751** | **1032** |
| **# Pulmonary**  **Hypertension (PH)** | **37** | **12** | **0** |
| **# Tetralogy of Fallot (TOF)** | **11** | **2** | **36** |
| **Ejection fraction (EF)**  **(SD) [%]** | **64.6 (5.6)** | **64.3 (6.0)** | **67.9 (3.1)** |
| **Diastolic vol.   (S.D.) [mL·m^-2^]** | **66.1(23.7)** | **64.9 (10.2)** | **____** |
| **Systolic vol.   (S.D.) [mL·m^-2^]** | **106.4 (13.5)** | **107.1 (13.5)** | **____** |
